# STIMULATE-ICP-Delphi (Symptoms, Trajectory, Inequalities and Management: Understanding Long-COVID to Address and Transform Existing Integrated Care Pathways Delphi): Study protocol

**DOI:** 10.1101/2022.04.06.22273514

**Authors:** Christina M. van der Feltz-Cornelis, Jennifer Sweetman, Gail Allsopp, Emily Attree, Michael G. Crooks, Daniel J Cuthbertson, Denise Forshaw, Mark Gabbay, Angela Green, Melissa Heightman, Toby Hillman, Lyth Hishmeh, Kamlesh Khunti, Gregory Y.H. Lip, Paula Lorgelly, Hugh Montgomery, W. David Strain, Emma Wall, Caroline Watkins, Nefyn Williams, Dan G. Wootton, Amitava Banerjee, the STIMULATE-ICP Consortium

## Abstract

**Introduction:** As mortality rates from COVID-19 disease fall, the high prevalence of long-term sequelae (Long COVID) is becoming increasingly widespread, challenging healthcare systems globally. Traditional pathways of care for Long Term Conditions (LTCs) have tended to be managed by disease-specific specialties, an approach that has been ineffective in delivering care for patients with multi-morbidity. The multi-system nature of Long COVID and its impact on physical and psychological health demands a more effective model of holistic, integrated care. The evolution of integrated care systems (ICSs) in the UK presents an important opportunity to explore areas of mutual benefit to LTC, multi-morbidity and Long COVID care. There may be benefits in comparing and contrasting ICPs for Long COVID with ICPs for other LTCs.

**Methods and analysis:** This study aims to evaluate health services requirements for ICPs for Long COVID and their applicability to other LTCs including multi-morbidity and the overlap with medically not yet explained symptoms (MNYES). The study will follow a Delphi design and involve an expert panel of stakeholders including people with lived experience, as well as clinicians with expertise in Long COVID and other LTCs. Study processes will include expert panel and moderator panel meetings, surveys, and interviews. The Delphi process is part of the overall STIMULATE-ICP programme, aimed at improving integrated care for people with Long COVID.

**Ethics and dissemination:** Ethical approval for this Delphi study has been obtained (Research Governance Board of the University of York) as have approvals for the other STIMULATE-ICP studies. Study outcomes are likely to inform policy for ICPs across LTCs. Results will be disseminated through scientific publication, conference presentation and communications with patients and stakeholders involved in care of other LTCs and Long COVID.

**Registration:** Researchregistry: https://www.researchregistry.com/browse-the-registry#home/registrationdetails/6246bfeeeaaed6001f08dadc/.

## Introduction

Despite major reductions in acute COVID-19 hospitalisation and mortality,[1] the persistence of symptoms over one year later is notable in the 45% of the 1.5 million individuals who had symptoms four weeks post-COVID in the UK.[2-4] Unlike some long-term conditions (LTCs), individuals with Long COVID (i.e. those with post-COVID symptoms >12 weeks) may still fully recover. However, new care pathways for Long COVID attempt to manage it akin to a LTC, given the increasing recognition of chronic symptoms.[4, 5]

Care pathways for LTCs have tended to be disease- or specialty-specific, an approach which fails to accommodate the heterogeneity of symptoms occurring in Long COVID. ICPs are structured, multi-disciplinary plans of the whole care pathway from primary care to specialist services and rehabilitation services, which may be better suited to Long COVID.[6-8] They offer coordination of investigation, treatment and rehabilitation, as well as opportunities for real-time iterative improvements in service design and delivery, quality and access to care, patient experience and satisfaction, while reducing complications and non-elective admission rates.[9-11] Evolution of integrated care systems (ICSs) in the UK provide opportunities to improve care across LTCs, multi-morbidity or multiple health conditions,[12] and Long COVID.

Long COVID encompasses a broad array of symptoms and symptom clusters. It is unlikely to reflect a single condition or pathology; rather it reflects a multi-faceted condition with numerous contributory factors: some identifiable, and others not yet understood.[13] Trajectory and recovery after SARS-CoV-2 infection are poorly defined and there is overlap with medically not yet explained symptoms (MNYES), referring to symptoms which do not represent a known medical condition, yet contribute significantly to lesser quality of life and treatment need.[14] Multi-organ complications,[13, 15-18] including neuropsychiatric sequelae (up to 20%), are well-documented.[19-24]

Table 1 shows current models that can potentially be applied to Long COVID ICPs. Depending on setting, expectations and provisions regarding treatment may differ. It can be argued that some current models for managing LTCs require improvement[25] as they cannot cover the whole range of patient presentations; episodic care is not appropriate for unpredictable exacerbating conditions, for example heart failure and COPD.[26, 27] An effort should be made to explore how to achieve integrated care from the perspective of individual conditions, but also from the perspective of how health services and settings can inform each other, and work together, to deliver optimal care for LTCs. The current pandemic and effort to set up Long COVID clinics[28] offers a unique opportunity to explore this from the perspective of Long COVID, and then to translate back to ICPs for other LTCs. For example, model 2 could also be based in primary care with better integration of GPs, primary care nurses and therapists. Model 3 could be more of a shared care arrangement between primary and secondary care with two-way data flow between these two sectors. In other words, one of the solutions for Long COVID care could be better working arrangements between community services, primary care and specialty care and ICSs might offer the perfect space for this in England.

**Table 1:** Potential models for ICPs managing recovery in Long COVID and other LTCs.

| Model | Example condition(s) | Recovery time | Managed by | Approach |
| --- | --- | --- | --- | --- |
| Model 1 | Community Acquired Pneumonia (CAP) | This may take 6 months to fully recover in terms of fatigue (NICE guideline)[29] | Primary care teams and community General Practitioners (GPs). There is a NHS CAP CQUIN aiming to support discharge from the hospital and safe follow up of these patients. Follow up imaging is usually arranged by secondary care. | Currently there is no well-developed integrated care pathway but there could be a chance to identify how to identify CAP follow up better based upon Long COVID care experiences. They support the patient through their recovery with the length of complete recovery and the ramifications for work often underestimated. |
| Model 2 | Post myocardial infarction, significant musculoskeletal injury | Taking a medium course to resolution, e.g., 1-2 years. | Multi-disciplinary team (MDT) driven and mostly provided in rehabilitation clinics. | Rehabilitation approach, personalised to the individual including a biopsychosocial approach to care, with physiotherapy and medical attention to address anxiety and depressive symptoms |
| Model 3 | A chronic disease like type 2 diabetes or stroke | It is managed but often recovery is not complete. | Usually managed in primary or community care, by GPs and diabetes nurses, or in the hospital setting. | Escalation of a small proportion with complex needs being managed in a specialist setting[30] |
| Model 4 | COPD Rheumatoid Arthritis | A chronic condition that may have high disability with tendency for relapses/exacerbations. | Limited care provision, mostly based in primary care with exacerbations increasingly managed in hospital in later stages. Growing emphasis on need to improve community diagnostics and | COPD is a condition that shares breathlessness as an important symptom with Long COVID, where pulmonary rehabilitation is a key evidence based treatment and supporting self-management is a key goal. Impact on function, breathlessness and psychological wellbeing as in Long COVID. Both conditions have a relapsing |
|  |  |  | where pulmonary rehabilitation is a key evidence-based treatment. | course of symptoms that may benefit from prompt intervention.<br>There is growing emphasis on the need to improve community diagnostics. |
| Model 5 | Comorbid mental disorders and other LTC.<br>For example, COPD in patients with mental disorders, often related to smoking.<br>Or, depression in diabetes patients. | These are in general chronic conditions with high disability.<br>There is an unmet clinical need here. | Mental disorders have case management, crisis teams, psychiatry follow up, but they do not identify physical health needs of their patients, such as respiratory issues. And clinics for somatic conditions can have short-term treatments available for psychological treatments but there is a lack of available long-term integrated treatment. | There are pilot playgrounds for dedicated respiratory clinics for patients with mental illness across the country.<br>Similar pilots exist for diabetes and depression - either community-based or hospital-based. |
| Model 6 | Encompassing multi-morbidity (i.e. more than two LTCs) as well as a spectrum of symptoms that do not fit into a usual pattern for diagnosis of a single disease i.e. MNYES,[14] or both, crossing the mental health and physical health divide. | The perceived burden of disease is high.<br>These are conditions requiring a multisystem approach. | No current consistent pathway of care exists. Consultation, collaborative care and decision aids supporting health care providers to provide ICP would be possibilities to link primary, community and specialist health care settings | These patients are highly likely to benefit from an ICP. This would be best served with a flag up system approach which is for people who don't quite meet full diagnostic criteria in one condition but almost meet it in many conditions. This would be labelled as MNYES but disease burden is high and there is a need to integrate physical and psychological health care provision. |

Acronyms: Community Acquired Pneumonia (CAP), National Institute for Health and Care Excellence (NICE), General Practitioner (GP), National Health Service (NHS), Commissioning for Quality and Innovation
(CQUIN), Multi-disciplinary team (MDT), Chronic Obstructive Pulmonary Disease (COPD), Long term condition (LTC), Integrated care pathway (ICP), Medically not yet explained symptoms (MNYES).

Even in well-defined entities such as community acquired pneumonia (CAP), symptoms such as fatigue[31] may take up to six months to resolve, even in young, physically fit people, fuelling fears that their symptoms will not abate, which may be biologically, psychologically, or socially driven, or depend on treatment setting. Learning from other LTCs, optimal Long COVID management may require a biopsychosocial model, rather than consideration of these components separately.[32]

## Research question

What are effective ICPs for individuals with Long COVID and how can they be transferred to other LTCs including mental disorders, multi-morbidity and the interface between known medical disorders and MNYES[14] as well as newly developed conditions?

## Materials and Methods

### Aims

In a Delphi study using a biopsychosocial approach, we will investigate:

1. (a) Key enabling elements for effective ICPs for individuals with long COVID, based on user experience, and (b) Strengths of existing ICPs for other LTCs.
2. Which (part of an) ICP model for Long COVID can be transferred to other LTCs for which care pathways were not yet developed sufficiently, and how.
3. Variations in uptake and adherence to treatment in Long COVID and LTCs.

### Study design and setting

This STIMULATE-ICP-DELPHI is a sub-study of the STIMULATE-ICP project (Symptoms, Trajectory, Inequalities and Management: Understanding Long COVID to Address and Transform Existing Integrated Care Pathways).[33] (**Figure 1**)

**Figure 1:**
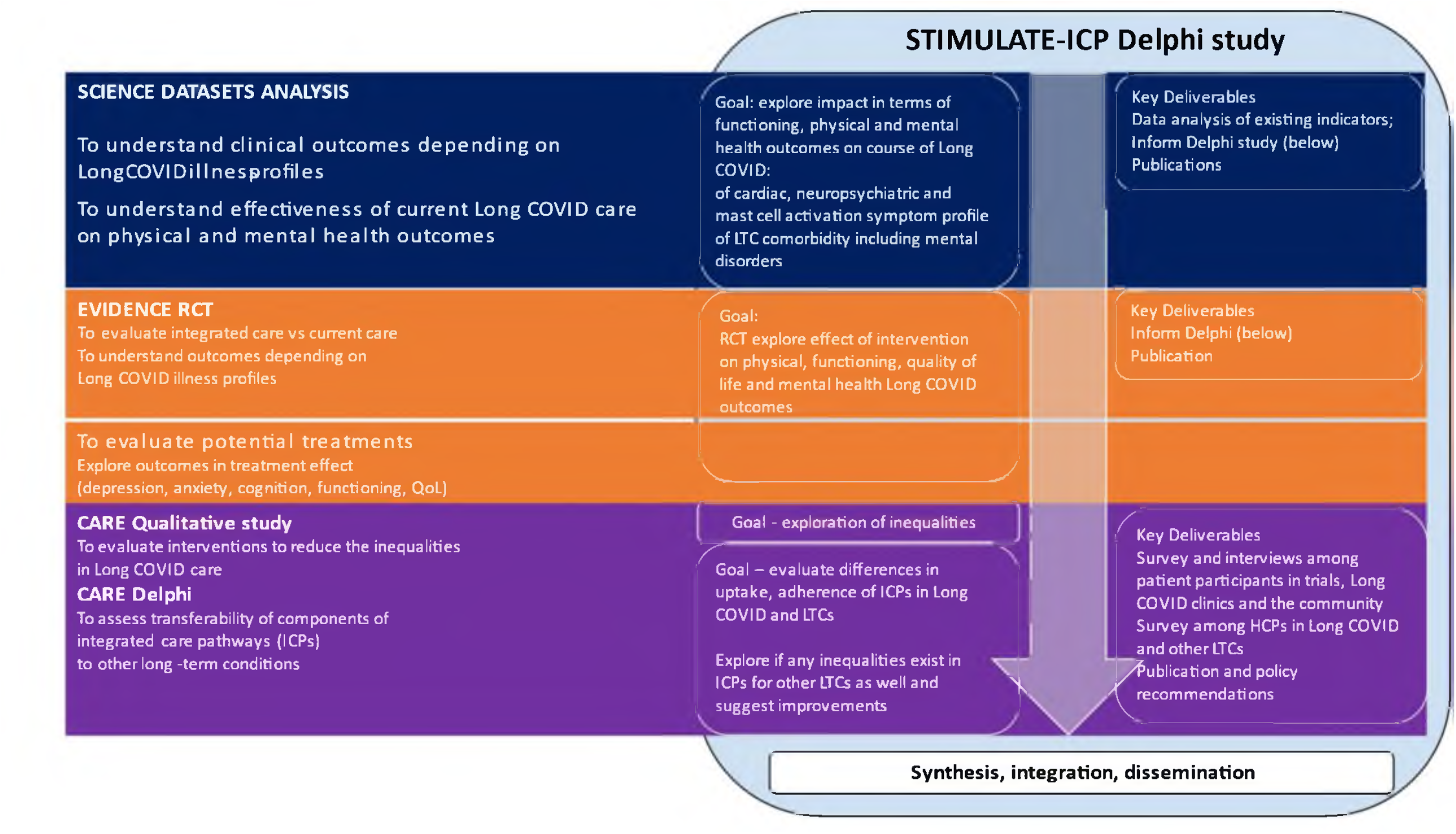
Flow chart showing the integration of STIMULATE-ICP DELPHI within overall STIMULATE-ICP

STIMULATE-ICP is funded by the National Institute for Health Research (NIHR: COV-LT2-0043)[34] and combines clinical epidemiological studies, a complex randomised trial exploring the benefit of an ICP for Long COVID (IRAS: 1004698), and mixed methods studies exploring inequalities of care and transferability of the ICP to other LTCs (IRAS: 303958).

The STIMULATE-ICP-DELPHI study will follow a Delphi process to establish consensus agreement on statements relating to ICPs and the transferability of ICP models between Long COVID and LTCs. The Delphi approach is a structured method for collecting opinions of experts concerning a subject of their expertise, reaching consensus over a number of rounds.[35] Since its development in the 1950s[36] a commonly used variation of the Delphi method is the *estimate-talk-estimate Delphi method* that combines assembling of expert opinions on an anonymous basis during surveys with open exchange during workshops moderated by a facilitator.[37] This Delphi method will be followed in this study,[38] aiming for stepwise consensus through three rounds of expert panel meetings involving exploration, prioritization, and as a final step attaining consensus(**Figure 2**).

**Figure 2:**
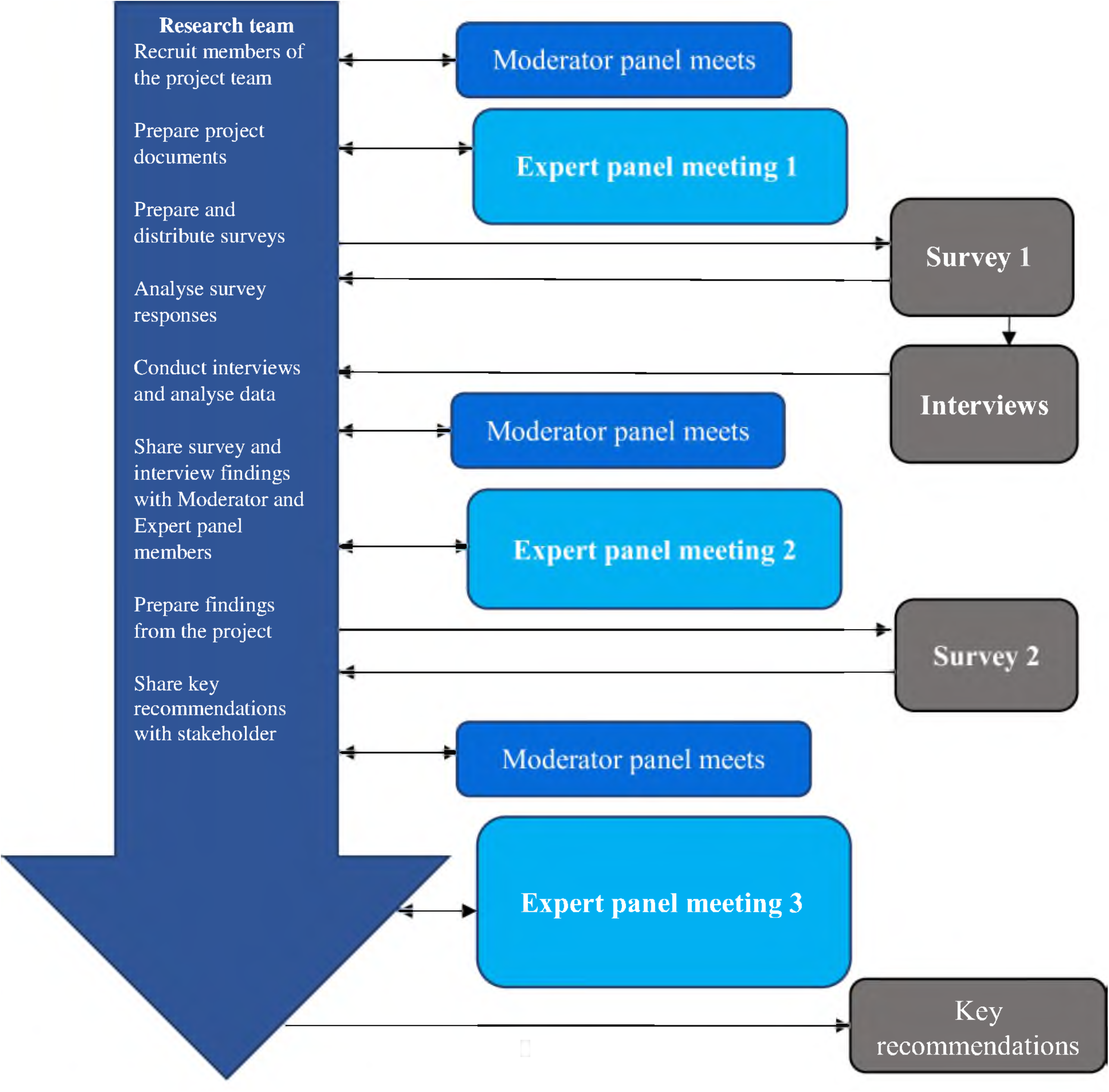
Outline of Delphi Process

The meetings will be interwoven by two online surveys, with addition of qualitative interviews. After a “knowledge check”, an information package based on the survey and interview outcomes will be provided to the panel. The expert panel will then discuss outcomes and provide policy recommendations in a final meeting.[39]

This study will depart from the theoretical framework of Goldberg and Huxley’s filter-model of access to care[40, 41] that describes four filters; three of which a patient has to navigate to enter a primary care treatment pathway, and a fourth to access specialist treatment. This model was originally developed for access to care for mental disorders, but it would be a good fit for exploring barriers and facilitators to entering Long COVID services and other LTCs not only for psychological symptoms but for physical symptoms as well. This extended model spans multiple healthcare challenges and extends the existing inequalities in health such as limited access to healthcare, incomplete pathways across community and hospital care, inadequate research translation to practice, and overall insufficient healthcare resources (**Figure 3**).

**Figure 3:**
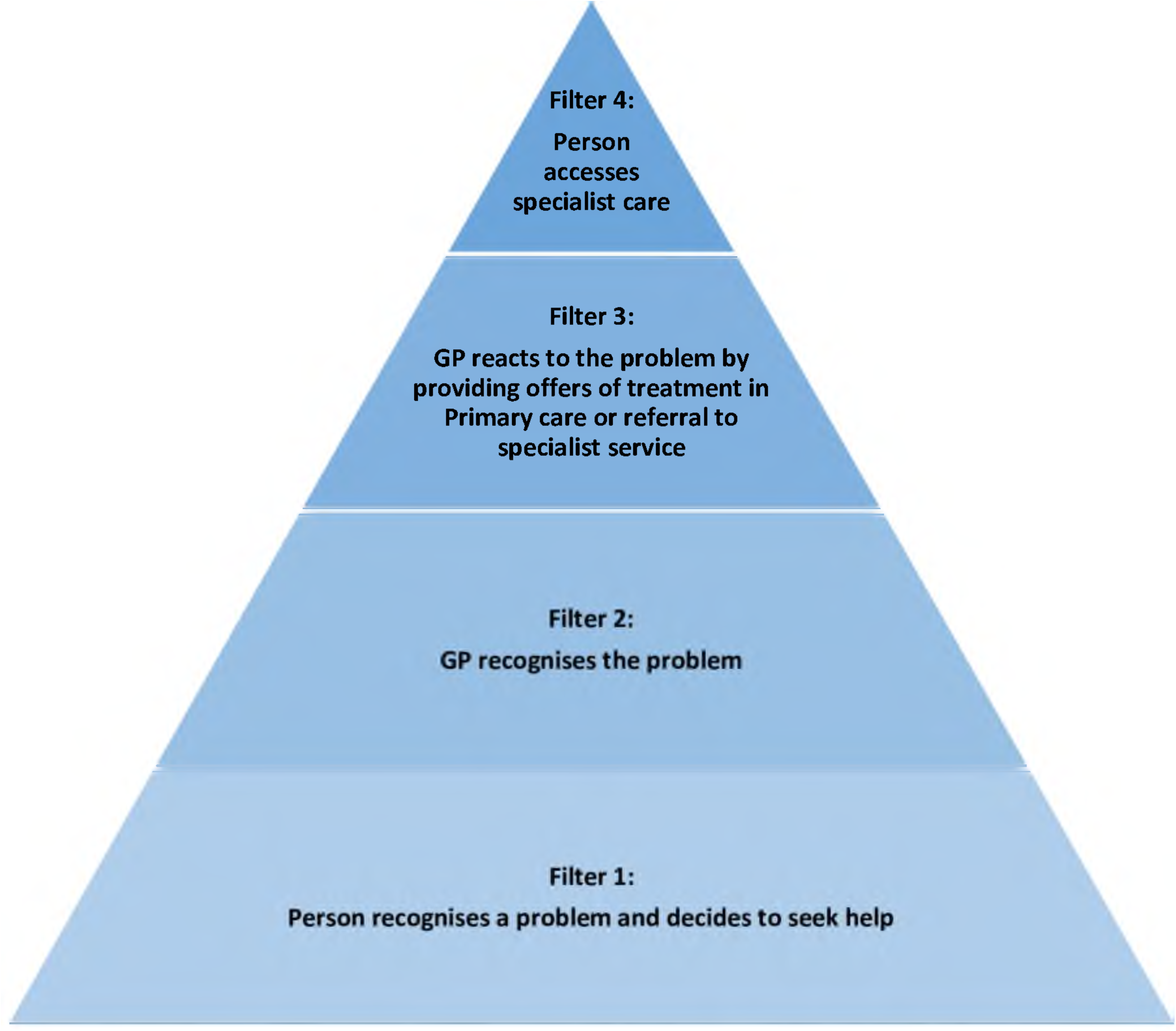
Filter model, expanded to conceptualise access to care for Long COVID or other long-term conditions (LTCs) including medically not yet explained symptoms (MNYES) and multi-morbidity

People with Long COVID and those with LTCs frequently present to healthcare services with multiple symptoms.[42-44] Therefore, this study will take account of competing demands to determine how people seeking support prioritise their symptoms when seeking help, and how healthcare providers deal with multiple symptoms reported when making decisions about appropriate care.[45, 46]

## Study management

This work will involve a research team (n=3), a moderator panel (MP) (n=3) and an expert panel (EP)(n=25). The research team will prepare, distribute and analyse all information for the project. The MP will meet regularly, providing advice and guidance to the research team to ensure scientific quality. The EP will include people with expertise in academic and clinical research in addition to lived experience of Long COVID, other LTCs such as cardiovascular disorders, type 2 diabetes, mental disorders and MNYES, or multi-morbidity; 5 of each group with a minimum of 25.

EP members will be selected by the MP building on suggestions from the Royal College of General Practitioners and national charities following a snowballing method. Patients with illness experience of Long COVID and other LTCs will be identified by clinics, medical trusts, patient networks and charities following a snowballing method. As part of the invitation process, the research team will inform prospective EP members about their role within the study. The EP will provide information and advice relating to their experiences of living with, or supporting people with Long COVID or LTCs.

### Sample size, participant characteristics, inclusion and exclusion criteria

Recruitment for Survey 1 will involve collecting quantitative and qualitative data from two purposive samples selected from community, primary care, and specialist settings. Sampling will seek to achieve sufficient numbers (minimum of N = 50 per group) in order to have a representative sample. Sample 1: patients/carers and clinicians with experience of Long COVID and other post COVID morbidity across England. Sample 2: patients/carers and clinicians involved in other LTCs. Participants in both samples will be recruited via the study website, with support from relevant online forums, associations and charities. As personalised care is now a specific NHS workstream which is intended to touch on all the LTC pathways, we will seek the opinion of ICS stakeholders (commissioners and those involved in the clinical networks) as to how personalised care should be delivered.[47-52]

The selection of interviewees for the qualitative study will be nested in the surveys. Participants from Survey 1, willing and able to provide informed consent, will be invited (using a separate survey link) to express interest in participating in an interview to discuss their experiences of healthcare relating to Long COVID and/or LTCs. Using details from the expression of interest form, the research team will select a purposive sample of people (minimum N=10-15) to interview. Within those volunteers, a maximum variation approach to participant selection will be adopted to ensure a wide range of experiences are reflected (accounting for experience of Long COVID and/or LTC(s), patient/clinician). Sampling will stop once saturation is reached. Both patients and clinicians will be interviewed as to how they deal with multi-morbidity or multiple symptoms and competing demands when accessing or providing healthcare, and the barriers and facilitators to providing or receiving ICP.[53-60]

### Data saturation

Interviews conducted for this research aim to supplement and provide context to the statements made by Survey 1 participants. For each group of interview participants (patients/clinicians with experience of Long COVID or LTCs), saturation will have been achieved when interviews stop providing new topics/themes which relate to ICPs.

### Patient and public involvement

The parent study STIMULATE-ICP has been enriched by robust patient and public involvement (PPI) using multiple channels, including regular updates and webinars, surveys, social media. The STIMULATE-ICP DELPHI study has been informed by existing engagements with people with experience of Long COVID/ LTCs in research, and PPI co-applicants who will contribute to methods and outputs. In addition, people with relevant disease experience will be involved in the EP and will be involved in the selection of other LTCs for comparison. Public and patients will be involved as stakeholders for this project, increasing awareness with relevant groups and promoting research activities. PPI will be involved in drafting the recommendations and their contributions through the EP and the wider STIMULATE-ICP team will shape our ultimate policy recommendations and the dissemination of this work. PPIE leads and co-applicants will contribute to the management and conduct of Delphi and qualitative interviews, the analysis plan and dissemination of the findings.

### Study processes

#### Round 1

During the initial EP meeting, the scope of this work will be agreed. This work will focus on adults (18 and over); outcomes will include confirming the list of relevant LTCs and those considered out of scope. As an inclusive approach, a variety of LTCs including mental disorders, multi-morbidity and the interface with MNYES will be considered. Fatigue, as a symptom, will be in scope, myalgic encephalomyelitis or chronic fatigue syndrome (ME/CFS) will be considered out of scope for this survey. Cognitive limitations will be in scope, non-capacity will be out of scope for the survey. Following confirmation of the scope, EP members will discuss and agree on questions to ask in Survey 1. The survey will include questions about:

1. Demographic factors (age, gender, ethnicity), relevant disease experience as patient or clinician, and clinical and work functioning profile of participants.
2. Experiences of Long COVID which prompted help-seeking with a focus on the process of seeking treatment, referrals, treatment(s) offered and received and whether there were any barriers or facilitators to that.
3. Challenges and advances for clinical care, knowledge gaps and policies, possible improvements to services, transferability of care models to other conditions will be explored for Long COVID and for other LTC ICPs.

#### Survey 1

This will be an anonymous, online survey (using the Qualtrics[61] platform) to establish demographic information and to explore the topics by open questions. Participants will also be invited to give a first indication of what their priorities to improve integrated care would be.

#### Round 1 Interviews

Qualitative semi-structured interviews will be used to examine the experience, and needs for treatment, of people living with Long COVID and other LTCs.

Examples of good practice will also be sought. Interview topic guides will be developed with guidance from the MP. Interviews will be offered over the telephone or a secure video-conferencing platform (zoom). Interviews are expected to last 40-50 minutes, but no longer than one hour, to reduce participant burden. With consent, they will be audio-recorded to allow verbatim transcription.[62-64] Where respondents appear fatigued, they will be given the option for the interview to take place over two time-periods, to have a family member present, or to shorten the length of the interview. The researcher will stop the interview at any point if participants indicate discomfort or distress.

#### Round 2

The EP will use data from Survey 1 and the interviews to create a list of statements about

1. Current Long-COVID clinics and future recommendations.
2. Current care models for LTCs.

Statements are likely to relate to how symptoms impact on general and social functioning and service use. Statements will also explore treatment and service need for people across different disease / condition profiles, in order to inform the recommendation phase.

#### Survey 2

There will be a second anonymous online survey seeking to explore for which statements consensus exists. Participants will review and respond to each of the statements using a 7-point scale (1. Totally disagree, 7, Totally agree).

#### Round 3

In a final meeting, the expert panel will use the findings to finalise a series of consensus-based recommendations about optimal care models for Long COVID and how these can be applied to other LTCs. These recommendations will be shared with healthcare professionals, policy makers and healthcare commissioners with the potential to influence future care.

### Data analysis and outcomes

Data analysis will provide descriptive statistics to outline the demographic characteristics of the two samples. Item response frequencies provide information about the current services offered. Open-ended questions will capture individual experiences of services and suggestions for future improvements. Responses will be organised into themes, with the research team adopting a pragmatic approach to provide feedback for the second expert panel meeting.

Thematic analysis will be conducted on data transcripts for round 1 interviews.[65] Theme development will be derived deductively from the topic guide. However, we will also allow for inductive theme development and will actively seek to identify new themes or topics within our data.[66] Following initial deductive and inductive coding, analysis will be set in the context of relevant theoretical concepts from the experience of chronic illness, such as, for example, the adapted Goldberg and Huxley’s filter model;[40] competing demands in primary care;[46] biographical disruption;[67] and illness careers.[68] Emergent patterns and early analysis will be discussed at regular research and moderator panel meetings for comment. Data from Survey 1 and Interviews will be combined and presented to the EP for consideration during round 2 of this study.

Data analysis will collate responses to Survey 2. Then consensus of opinion about each statement will be assessed using interquartile deviations (IQD). For this calculation, at least 50% of individuals will have responded using the same category. IQD ≤ 1 is considered to indicate consensus. Findings from survey 2 will be combined with an information pack based upon input from other STIMULATE-ICP sub studies as lined out in Figure 1, and shared with the EP members.

### Data management plan

This study will produce online survey data and qualitative interview data. Online surveys will be anonymous and therefore a survey ID code will be created for participants (for example, S146 would be the code given to Survey 1 participant number 46). Data will be downloaded from Qualtrics to Microsoft Excel. Qualitative interview data will be audio recorded via Zoom (for virtual interviews or telephone interviews). Participant ID codes will be provided to all participants (for example, DI07 would be the code given to Delphi Interview participant number 7). A password-protected Microsoft Excel file will be used to track the status of data preparation for each interview; this document will contain participant names and ID codes. Audio recording will be transcribed verbatim into Microsoft Word documents. Transcripts will then be anonymised ready for analysis.

All data will be stored electronically on the University of York secure server with access restricted to the research team involved with this project. Analysis will be conducted in Microsoft Excel, SPSS and NVivo. Anonymous data (such as Survey 1 original responses) will be shared with MP and EP members to enable discussions and decisions about the organisation of data and the development of statements.

### Ethics

This Delphi study was reviewed and approved by the University of York Department of Health Sciences Research Governance Committee in December 2021 (HSRGC/2021/478/A:STIMULATE).

### Informed consent

#### Survey

Regarding the survey, following the presentation of participant information, consent for anonymous data to be collected, analysed and disseminated as part of this project will be required before survey questions are displayed for Surveys 1 and 2.

#### Interview

Regarding the interview, a separate survey link will be embedded at the end of Survey 1 to enable participants to express an interest in participating in a subsequent qualitative interview without linkage to their survey answers. All individuals who express an interest in being interviewed will be contacted to confirm whether they have been selected to contribute to the interviews. Individuals who are selected for interview will then receive full interview study information and will be required to provide consent if they wish to participate in an interview.

#### Data handling

The study is compliant with the requirements of General Data Protection Regulation (2016/679) and the UK Data Protection Act (2018). All investigators involved in the study will comply with the requirements of the General Data Protection Regulation (2016/679) with regards to the collection, storage, processing and disclosure of personal information, and will uphold the Act’s core principles. For this STIMULATE-ICP Delphi sub-study, survey data will be downloaded and stored/archived at the University of York. All interviews will be recorded. Interview data will be transcribed and coded by JS and will be identified and stored/archived at the University of York. Information provided to survey and interview participants will outline their right to withdraw at any point during this research. Data collected up to the point of withdrawal will be used unless there is an expressed request for withdrawal of all data.

#### Safety considerations

There are not considered to be any safety concerns for participants involved with this project. EP and MP members will be informed of the project aims, the focus of their role and the project timescales prior to joining the study. These are voluntary roles and individuals can withdraw from the study at any time. Likewise, Survey participants’ data will be shared anonymously with basic demographic details being collected to enable researchers to describe the sample. Survey participants will be able to ask questions to the research team, provide consent and withdraw at any point. Interview participants will provide contact details to the research team to enable interviews to be organised and conducted. Transcribed data will be anonymised, and interview participants can stop or pause interviews at any point should they wish to. All data collected will be online, virtually or using the telephone to minimise any burden for participants. The anticipated time to complete each research activity will be shared with potential participants to enable them to make informed decisions about whether to participate in each element of the study.

### Status and study timeline

Jan-Feb 2022 – Recruitment of Expert Panel members

March 2022 – Initial Expert Panel meeting

April 2022 – Survey 1 launched online

May 2022 – Interviews started

June 2022 – Survey 1 closed

July 2022 – Interviews completed, Survey 1 data cleaning and analysis

August 2022 – Survey 1 data analysis, Interview data transcription, small groups of Expert Panel members discuss preliminary organisation of data from Survey 1

September 2022 – Second Expert panel meeting, Interview data analysis

October 2022 – Launch of Survey 2 online, Interview data analysis

November 2022 – Interview data analysis

December 2022 – Survey 2 closed, Interview data analysis

January 2023 – Survey 2 data cleaning and analysis

Feb/March 2023 – Final Expert Panel meeting

March 2023 – Key recommendations finalised and disseminated

## Discussion

### Dissemination

We will publish the findings from this Delphi study in peer reviewed journals and will present the findings during conferences. Table 2 provides an overview of the proposed deliverables for stakeholders during the study.

**Table 2.** Dissemination to stakeholders.

|  |  |  |  |
| --- | --- | --- | --- |
| Early deliverables | Early read out and contact with stakeholders (NHS England, the Royal College of GPs, AHSNs, other Royal Colleges, RCP, RCPsych, patient groups and NICE) |  |  |
| What should integrated care look like? What should be integrated for Long COVID, other LTCs and multimorbidity? |  |  |  |
| How can ICPs integrate primary and specialty health care, as well as somatic and mental health care? |  |  |  |
| Patient and clinician experiences in ICPs and variety in Long COVID and other LTCs regarding funding and structure of their care pathways. |  |  |  |
| Inform care pathways for other LTCs that are unheard or under-served. |  |  |  |
| What are facilitators and barriers to integrated care pathways? |  |  |  |
| Final deliverable | Contact with stakeholders (NHS England, the Royal College of GPs, AHSNs, other Royal Colleges, RCP, RCPsych, patient groups and NICE) |  |  |
| Outputs | Webinar contact with stakeholders | Report with summary of findings for stakeholders | Policy recommendations |
| Novel models of provision across sectors, addressing biological, psychological and social components as a matter of routine, new multi-disciplinary groups in settings such as primary care. |  |  |  |

### Conclusions

The pandemic and the legacy of Long COVID will alter the landscape of the UK NHS forever, and possibly health care systems in other countries as well. This Delphi study can support a novel way of developing integrated models of care. It will inform the beginning of a change in NHS integrated care systems across diseases and the primary and specialty health care divide, while putting the patient first.

## Data Availability

This is a protocol paper; no data are available from this work at present.

## Authors’ Contributions

Conceptualisation and methodology: CFC, JS, GA, EA, MG, AB, STIMULATE-ICP team. Funding acquisition: all authors. Ethics: CFC, AB, PM. Project administration: CFC. Original manuscript drafting and preparation: CFC, JS. Review and editing of manuscript: all authors. Patient and public involvement: LH, EA. All authors approved the final version of the article.

## Acknowledgements

This paper has been published on behalf of the STIMULATE-ICP Consortium: Professor Amitava Banerjee (Chief Investigator), Professor Paula Lorgelly, Professor Elizabeth Murray, Dr Hakim-Moulay Dehbi, Professor Hugh Montgomery, Dr Yi Mu, Sarah Clegg, Dr Mel Ramasawmy–University College London; Dr Melissa Heightman (Co-Chief Investigator), Dr Toby Hillman, Dr Emma Wall, Dr Michael Zandi—University College London Hospitals NHS Foundation Trust; Professor Dame Caroline Watkins, Denise Forshaw, Dr Gordon Prescott–University of Central Lancashire; Dr Gail Allsopp— Derbyshire NHS Foundation Trust/Royal College of General Practitioners; Professor Mark Gabbay, Professor Gregory Lip, Professor Dan Cuthbertson, Dr Dan Wootton, Professor Nefyn Williams—University of Liverpool; Dr Michael Crooks—University of Hull; Dr Angela Green—Hull University Teaching Hospitals Trust; Professor Christina van der Feltz-Cornelis, Dr Jennifer Sweetman, Dr Han-I Wang, Natalie Smith—University of York; Professor Kamlesh Khunti—University of Leicester; Dr David Strain—University of Exeter; Dr Emily Attree, Jasmine Hayer, Rachel Hext, Lyth Hishmeh, Kim Horstmanshof, Mag Leahy, Antony Loveless, Clare Loveless, Rita Mallinson Cookson, Andrew Williams, Rachel Williams–PPI Representatives; Dr Nisreen Alwan, Dr Donna Clutterbuck—University of Southampton; Dr Marija Pantelic—University of Sussex; Chris Robson—Living With COVID Recovery; Professor Sir Mike Brady, Dr Rajarshi Banerjee, Dr Cat Kelly– Perspectum. An up-to-date version of Consortium members can be found: https://www.stimulate-icp.org/team. STIMULATE-ICP can be contacted at: Papiya Mazumdar contributed to obtaining ethics approval.

